# The prevalence of hepatitis C virus (HCV) infection in *β*-thalassemia patients in Pakistan: a systematic review and meta-analysis

**DOI:** 10.1101/19011973

**Authors:** Sohail Akhtar, Jamal Abdul Nasir, Farrukh Shah, Andrew Hinde

**Affiliations:** Department of Statistics, Government College University Lahore, Lahore, Pakistan; Consultant haematologist, Department of haematology Whittington Health Magdala Avenue London, UK; Department of Social Statistics and Demography, School of Economic, Social and Political Sciences, University of Southampton, SOUTHAMPTON, UK

**Keywords:** Prevalence, HCV, *β*-thalassemia patients, Pakistan, systematic review and meta-analysis

## Abstract

**Objective:** Hepatitis C virus (HCV) infection is the most commonly reported bloodborne infection in Pakistan. Frequent blood transfusions in *β*-thalassemia patients expose them to a high risk of HCV infection. The purpose of this paper is to summarize the current data on the prevalence of HCV infection among *β*-thalassemia patients in Pakistan by using a systematic review and meta–analysis.

**Design:** Systematic review and meta-analysis.

**Participants:** *β*-thalassemia patients in Pakistan.

**Data sources:** Following PRISMA guidelines, a comprehensive literature search in PubMed/MEDLINE and EMBASE was performed to identify published articles reporting on the prevalence of HCV among *β*-thalassemia patients in Pakistan. Only English language articles were considered. Two independent authors selected studies. The methodological quality of the included studies was assessed using the Quality Assessment Tool for Observational Cohort and Cross-Sectional Studies.

**Results:** The search conceded a total of 138 studies, of which 27 studies were finally considered for meta-analysis. The pooled prevalence of HCV in *β*-thalassemia patients in Pakistan was 36.21% (95% CI: 28.98– 43.75%) based on 5,789 *β*-thalassemia patients, but there was considerable heterogeneity. Meta-analysis estimated the HCV prevalence among the *β*-thalassemia patients at 45.98 % (95% CI: 38.15–53.90%) in Punjab, 31.81% (95% CI: 20.27–44.59%) in Sindh, and 28.04% (95% CI: 13.58–45.26%) in Khyber Pakhtunkhwa. Meta–regression analysis showed that geographical location was a key source of heterogeneity.

**Conclusions:** The pooled prevalence of hepatitis C virus among *β*-thalassemia patients in Pakistan was 36.21%, but varies regionally within the country. The prevalence is higher than in neighboring countries. With the use of standard prevention procedures during blood transfusion, the risk of HCV transmission among *β*-thalassemia patients could be controlled hence prevalence of HCV in *β*-thalassemia patients could be reduced.

**Strengths and limitations of this study:**

➢ This is the first systematic review and meta-analysis to estimate the pooled prevalence of HCV infection among the *β*-thalassemia patients in Pakistan
➢ We used of an extensive search strategy and adherence to predetermined inclusion and exclusion criteria.
➢ Strong and reliable methodological and statistical methods were used.
➢ Our analyses possessed a considerable amount of quantifiable heterogeneity.
➢ Not all regions in Pakistan were represented and most of the included studies were hospital-based, making it difficult to generalize the findings of this review.

## Introduction

The *β*-thalassemias are among the most common genetic diseases and affect millions of children throughout the world.^1^ Around 1.5% (80-90 million people) of the global population are carriers for *β*-thalassaemia, with 50,000-60,000 new *β*-thalassemia patients being born each year.^2^ *β*-thalassemia is most prevalent in the populations of Asia, the Indian subcontinent, the Mediterranean region, Africa and the Middle East.^3-5^ In Pakistan, *β*-thalassemia is one of the commonest inherited disorders, with a carrier frequency of 5% to 7% in the Pakistani population.^2^ *β*-thalassemia patients are now surviving to older ages due to the availability of blood transfusion and iron chelation. There are around 100,000 patients registered currently but the burden of disease is increasing, with 5,000 to 9,000 children born with the disorder annually.^6^ Bloodborne infections are the second commonest cause of death in *β*-thalassemia patients in Pakistan.^2^ Patients with *β*-thalassemia are at high risk of developing hepatitis C (HCV) infection due to regular blood transfusions, especially if adequate viral screening of blood donors has not been undertaken. The infection risk in *β*-thalassemia patients acts as a marker for the risk of transfusion transmitted infections in the general population as their exposure to blood transfusions is high. If the infection rate is low in *β*-thalassemia patients it means that the risk for the general population will be minimal.

Hepatitis C virus is one of the most common bloodborne viruses. More than 10 million people are living with Hepatitis C virus (HCV) in Pakistan with its associated high morbidity and mortality.^7^ Pakistan is a developing country, and according to the human development index of the United Nations, it stands in 150th position out of 189 countries and territories.^8^ The health standard in Pakistan is below the international level. Therefore, transfusion of contaminated blood is still a major risk factor for the spread of hepatitis C. This is due to the lack of appropriate donor screening and the widespread use of paid blood donors.^9^ Several studies have been reported on the prevalence of HCV among *β*-thalassemia patients in Pakistan and there is considerable variation in the prevalence reported in the individually published studies. The purpose of this systematic review and meta-analysis is to identify, select, summarize and estimate the pooled prevalence based on the available published studies conducted on the prevalence of HCV infection among *β*-thalassemia patients, and its associated risk factors in Pakistan. To the best of our knowledge, this is the first systematic review and meta-analysis to estimate the pooled prevalence of HCV infection among *β*-thalassemia patients in the country.

## Methods

### Search strategy

A comprehensive literature search on Medline, PubMed, EMBASE, the Cochrane Library, and Pakistani Journals Online websites was conducted to identify studies performed on the prevalence of HCV infection among *β*-thalassemia patients and published up to 31 May 2019. Using MeSH headings, the terms “prevalence”, “epidemiology”, “seroprevalence”, ‘‘hepatitis C Virus’’, “HCV”, “hepacivirus”, ‘‘hep C,’’ ‘‘thalassemia,’’,” *β*-thalassemia”, “thalassemia major” “multitransfused blood transfusion”, “patients”, “Pakistani”, and ‘‘Pakistan” as well as variations thereof were searched for. The results were defined using the Preferred Reporting Items for Systematic and Meta-analyses (PRISMA) guidelines (Table 1)^10^, and the PRISMA 2009 Checklist is attached in supplementary file S1.

**Table 1.** Description and list of characteristics of included studies

| Author | Year | Study Design | Sample size | Cases | Prevalence (%) | Setting | Province | Sex | Working Year | % Female | % Male | Average Age | Test | Quality |
| --- | --- | --- | --- | --- | --- | --- | --- | --- | --- | --- | --- | --- | --- | --- |
| Bhatti et al.<sup>18</sup> | 1995 | NA | 35 | 21 | 60.00 | Urban | Khyber Pakhtunkhwa | Both | NA | 14.28 | 85.71 | 6.5 | ELISA | Medium |
| Muhammad et al.<sup>19</sup> | 2003 | Cross-sectional | 80 | 29 | 36.25 | Urban | Khyber Pakhtunkhwa | Both | Jul. 1999 to Mar. 2001 | NA | Na | 7.5 | ELISA | Medium |
| Shah et al.<sup>20</sup> | 2005 | Cross-sectional | 250 | 142 | 56.80 | Urban | Khyber Pakhtunkhwa | both | Jan. 2000 to Jan. 2001 | 72.00 | 28 | 10 | ELISA | Medium |
| Hussain<sup>21</sup> | 2008 | Cross-sectional | 180 | 75 | 41.67 | Urban | Khyber Pakhtunkhwa | Both | Jan. 2002 to Dec. 2003 | NA | NA | 6.8 | ELISA | Good |
| Ali et al.<sup>22</sup> | 2011 | NA | 167 | 26 | 15.57 | Urban | Khyber Pakhtunkhwa | Both | NA | 62.28 | 36.7 | NA | RNA | Medium |
| Khattak et al.<sup>23</sup> | 2013 | NA | 170 | 37 | 21.67 | Urban | Khyber Pakhtunkhwa | both | Jan. 2012 to Dec. 2012 | 55.29 | 44.71 | 10 | ELISA | Medium |
| Khan et al.<sup>24</sup> | 2015 | Cross-sectional | 180 | 14 | 7.77 | Urban | Khyber Pakhtunkhwa | Both | Jun 2013 to Jul 2014 | 38.89 | 61.11 | NA | NA | Medium |
| Shah et al.<sup>25</sup> | 2018 | NA | 324 | 18 | 5.56 | Urban | Khyber Pakhtunkhwa | Both | Oct. 2013 to Mar. 2014 | 34.50 | 60.23 | 15.5 | RNA | Medium |
| Younus et al.<sup>26</sup> | 2004 | Cross-sectional | 75 | 32 | 42.00 | Urban | Punjab | Both | Jul to Sept 2003 | 64.00 | 36 | 6.5 | ELISA | Good |
| Iqbal et al.<sup>27</sup> | 2010 | NA | 141 | 50 | 35.46 | Urban | Punjab | both | Sep. 08 to Aug. 09 | 58.20 | 41.8 | 8 | ELISA | Medium |
| Ain et al.<sup>28</sup> | 2011 | Cross-sectional | 300 | 195 | 65.00 | Urban | Punjab | Both | NA | 34.33 | 65.67 | 10 | NA | Medium |
| Iqbal et al.<sup>29</sup> | 2013 | Cross-sectional | 95 | 40 | 42.11 | Urban | Punjab | both | Oct. 2009 Apr. 2010 | 60.00 | 40 | 9.2 | ELISA | Medium |
| Din et al.<sup>30</sup> | 2014 | NA | 95 | 45 | 47.00 | Both | Punjab | Both | Jul. to Sept. 2017 | 56.84 | 53.68 | 7 | ELISA | Good |
| Nazir et al.<sup>31</sup> | 2014 | NA | 200 | 82 | 41.00 | Urban | Punjab | Both | Jan. to May 2013 | 12.00 | 88 | 8.5 | ELISA | Medium |
| Saeed et al.<sup>32</sup> | 2015 | Cross-sectional | 262 | 146 | 55.73 | Urban | Punjab | Both | Nov., 2011 to April 2012 | 40.07 | 59.92 | 9.26 | ELISA | Medium |
| Sheikh et al.<sup>33</sup> | 2015 | Cross-sectional | 145 | 99 | 68.27 | Urban | Punjab | Both | Jan 2009 to Dec | 63.45 | 36.55 | 9 | ELISA | Medium |
| Khan et al.<sup>34</sup> | 2017 | Cross-sectional | 470 | 216 | 45.95 | Urban | Punjab | Both | Mar. 2014 to Sept. 2014. | 65.96 | 34.04 | 4.8 | ELISA | Medium |
| Rashid et al.<sup>35</sup> | 2017 | Cross-sectional | 130 | 27 | 20.76 | Urban | Punjab | Both | Jan. 14 to Jun 14 | 60.00 | 40 | 9.7 | ELISA | Medium |
| Raza et al.<sup>36</sup> | 2018 | Cross-sectional | 200 | 82 | 41.00 | Urban | Punjab | Both | Jan. 2015 to Dec. 2016 | 43.00 | 57 | 10.11 | ELISA | Good |
| Mujeeb et al.<sup>37</sup> | 1997 | NA | 91 | 46 | 50.54 | Urban | Sindh | Both | NA | 39.56 | 60.43 | 13 | NA | Medium |
| Akhtar et al.<sup>38</sup> | 2002 | Cross-sectional | 341 | 70 | 20.50 | Urban | Sindh | Both | Jun-91 | NA | NA | 5 | RNA | Good |
| Akhtar et al.<sup>39</sup> | 2004 | NA | 86 | 38 | 44.20 | Urban | Sindh | Both | NA | 31.40 | 67.44 | 12 | ELISA | Medium |
| Riaz et al.<sup>40</sup> | 2011 | Cross-sectional | 79 | 34 | 43.00 | Urban | Sindh | Both | Jul-Sept. 2009 | 41.77 | 58.23 | 12 | ELISA | Medium |
| Ansari et al.<sup>41</sup> | 2012 | Cross-sectional | 160 | 21 | 13.10 | Urban | Sindh | Both | Jan. to Dec. 2010 | 49.38 | 50.63 | 8.5 | ELISA | Medium |
| Sultan et al.<sup>42</sup> | 2016 | Cross-sectional | 100 | 27 | 27.00 | Urban | Sindh | Both | Jun 2011 to Jun 2014 | 54.00 | 46 | 15 | ELIZA | Good |
| Burki et al.<sup>43</sup> | 2005 | NA | 180 | 75 | 41.67 | Urban | Punjab + Khyber Pakhtunkhwa | both | Jan. 2002 to Dec. 2003 | NA | NA | 6 | ELISA | Medium |
| Kiani et al.<sup>44</sup> | 2016 | Cross-sectional | 1253 | 273 | 21.71 | Urban | Punjab + Sindh | Both | Jul. to Dec. 2015 | 46.21 | 53.79 | 10.1 | NA | Medium |

### Inclusion and exclusion criteria

Studies were included in the meta-analysis if: 1) Only studies published in peer-review journals. 2) Studies conducted in Pakistan. 3) Studies reporting on the prevalence HCV in thalassemia patients. 4) Articles published in the English language.

Studies were excluded if: 1) Studies in non-English languages. 2) Case series, reviews, letters, and editorials or commentaries. 3) if we cannot find the prevalence from the article data 4) Duplicates (overlapped data); for studies published in more than one articles, the up-to-date version was considered. 5) Pakistani community living outside Pakistan.

### Data extraction

After choosing the relevant articles, two reviewers (J.A.N and S.A.) independently screened the titles and abstracts to consider articles for full-text review, and extracted all the necessary data using a standardized data extraction format of Microsoft Office Excel 2013. The extracted information was: surname of first author, year of study, year of publication, geographic region (province), gender, study design, study setting (rural, urban or both), sample size and average age of *β*-thalassemia patients. Any disagreement regarding the extracted information was resolved by discussion and mutual consensus.

### Evaluating the quality of the included studies

Two authors (J.A.N. and S.A.) also independently judged the methodological quality of each included study using Quality Assessment Tool for Observational Cohort and Cross-Sectional Studies.^11^ Any disagreement on the quality assessment check list was resolved by discussion and consensus. We categorized the quality of each included study as good if its points was at least 70%, medium if its points were lying between 50%-69%, and poor if its points was less than 50%.

### Statistical analyses

Statistical analyses were performed using the software R version 3.5.3.^12^, using two packages: meta 4.9-2 and metafor 2-0. Random effects (DerSimonian and Laird) models were used to make point estimates and their 95% confidence intervals (95% CI), as well as to estimate the pooled prevalence of HCV among the *β*-thalassemia patients. A process for combining prevalence in the meta-analysis of multiple studies was used and the results presented in a forest plot. Random effect models are more conservative than the fixed effect model, and have better properties in the presence of heterogeneity, as the random effect model allows both within-study and between-study variances.^13-14^ The Freeman–Tukey Double Arcsine transformation was used to stabilize the variance prior to the calculation of the pooled estimates.^15^ Heterogeneity among the eligible articles was investigated with the *I*^2^ index.^16^ The heterogeneity (I^2^-index) is categorized as low (25%), moderate (50%) and high (75%). To determine the possible reasons for substantial heterogeneity, univariable meta-regression and subgroup analyses were conducted by geographical location, sample size, year of publication, year of data collection, gender and average age of the *β*-thalassemia patients. The existence of publication bias was initially assessed by visually inspecting a funnel plot and then tested using Egger’s test.^17^

## Results

### Literature search

Initially, 138 potential studies were identified. Of these, 35 were duplicates and were removed. The remaining 103 studies were screened by title and abstract. After reading the titles and abstracts, 62 studies were deemed irrelevant and excluded from the meta-analysis. As a result, only 41 articles were selected for full text reading. For the following reasons 14 studies were excluded after full text read: articles with no numerical prevalence measure(s) of HCV in *β*-thalassemia patients; studies that were not based in Pakistan; studies that provided combined HCV and hepatitis B virus prevalence; studies based on duplicated data sets or that did not meet the eligibility criteria or failed to include relevant indicators. In the end, 27 studies met the inclusion criteria and data were extracted for the analysis. The PRISMA flow diagram of study selection process is presented in Fig. 1^10^.

**Figure 1:**
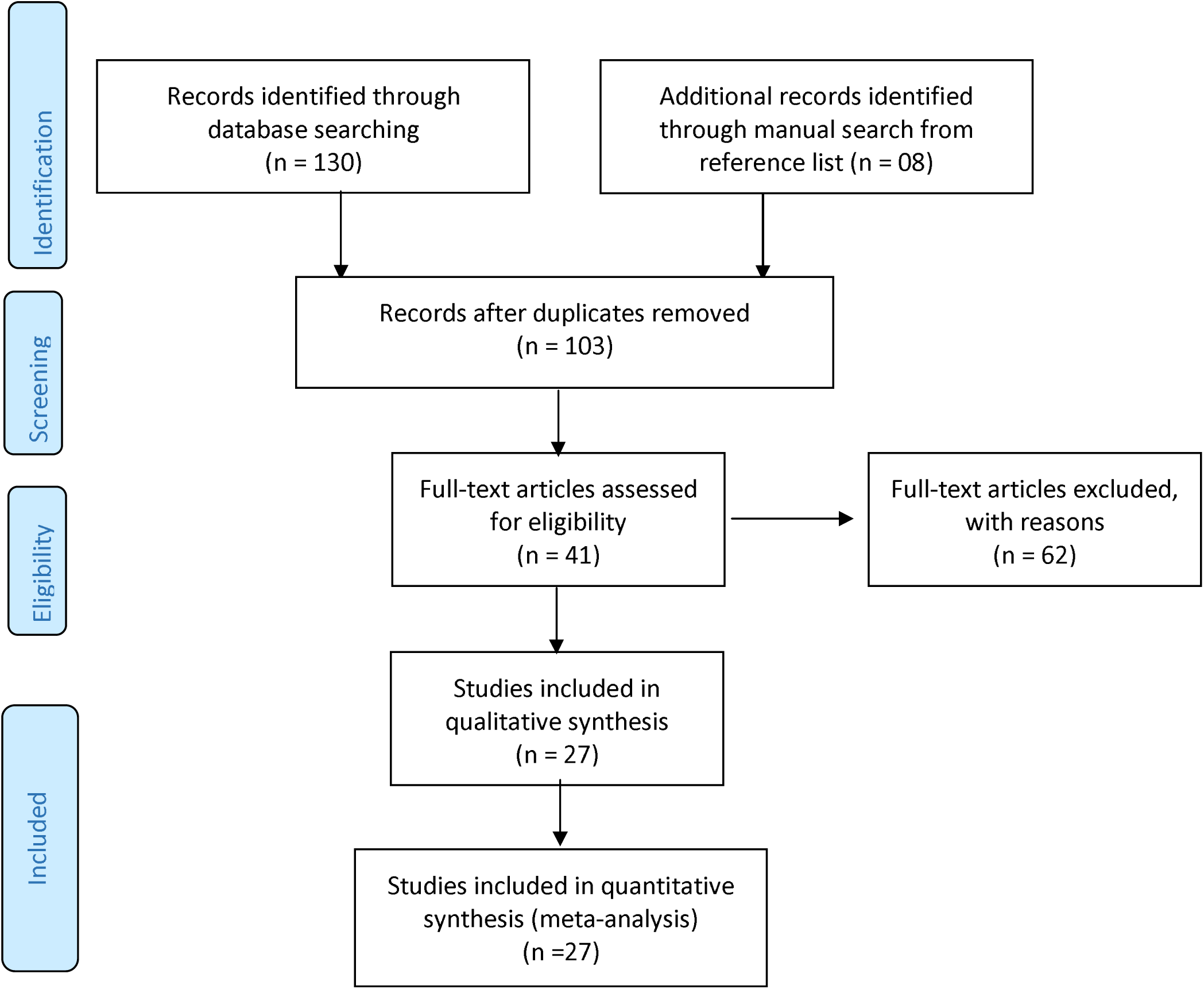
PRISMA 2009 flow diagram^10^ explaining the number of included and excluded articles in the meta-analysis on the prevalence of HCV in *β*-thalassaemia patients in Pakistan

### Characteristics of included studies

The main characteristics of the selected articles are summarized in Table 1. A cross-sectional research design was used in 17 studies, whereas ten studies did not clearly specify the research design. The articles were published between January 1995 and December 2018 while the period of subject inclusion was from June 1991 to September 2017. Three provinces of Pakistan were represented in the included studies: eight were conducted in Khyber Pakhtunkhwa,^18-25^ 11 were conducted in Punjab^26-36^ and six were conducted in Sindh.^37-42^ One study was conducted both in Punjab and Khyber Pakhtunkhwa^43^ while one was conducted in Punjab and Sindh.^44^ Most of the included studies (20 out of 27) reported HCV prevalence based on the results of the ELISA (enzyme-linked immunosorbent assay) test.^17-20, 22, 25, 26, 28-35, 40-43^ Only three studies reported the confirmation of HCV infection by RNA test.^21, 24, 39^ Four studies did not report the type of assay used for HCV antibody reactivity testing.^23, 27, 36, 38^ The sex of the patients was reported in 23 studies. The proportion of females ranged from 28.0% to 88.0%. The average age of patients varied from 4 years^34^ to 15.5 years^25^. After reviewing the quality of the studies, six were deemed to be of good quality, 21 of moderate quality, and no article was found with poor quality. Sample size varied among studies with the smallest having a total of 35 patients^17^ and the largest 1,253 patients.^35^

### Prevalence of HCV in β-thalassemia patients

Table 2 shows the summary of statistical analyses of the prevalence of the HCV among *β*-thalassemia patients in Pakistan. The overall prevalence of HCV infection among *β*-thalassemia patients was 36.21% (95% CI: 28.98– 43.75%, *I*^2^ = 97.0%; 27 studies), based on a pooled sample of 5,789. Forest plot of HCV prevalence among the *β*-thalassemia patients in the three provinces of Pakistan is presented in Fig. 2. The funnel plot (Fig. 3) visually showed no publication bias and supported by the results of Egger’s test (p = 0.1506). Table 2 also presents the prevalence of HCV among *β*-thalassemia patients for subgroups. The pooled subgroup prevalence stratified by geographical location (province) revealed that the prevalence of HCV among *β*-thalassemia patients were high 45.98% (95% CI: 38.15-53.90%; *I*^2^ = 92.3%, based on 11 studies) in Punjab, compared with 31.81% (95% CI: 20.27-44.59%; *I*^2^ = 92.8%; based on 6 studies) of Sindh and 28.04% (95% CI: 13.58-45.26%, *I*^2^ = 97.6%; based on 8 studies) of Khyber Pakhtunkhwa. There was no significant difference between the prevalence of HCV among males (34.71% (95% CI: 23.32-47.04%)) and female (32.31% (95% CI: 20.17-45.75%)) *β*-thalassemia patients. The prevalence of HCV among *β*-thalassemia patients increased significantly with age-group: the prevalence among *β*-thalassemia patients below 10 years of age was 33.87% (95% CI: 18.93-50.62%, *I*^2^ = 96.2 %; 9 studies) while above 10 years age-group 51.51% (95% CI: 34.52-68.34%, *I*^2^ = 96.2%; 9 studies). There was no publication bias for all subgroup analyses.

**Table 2.**
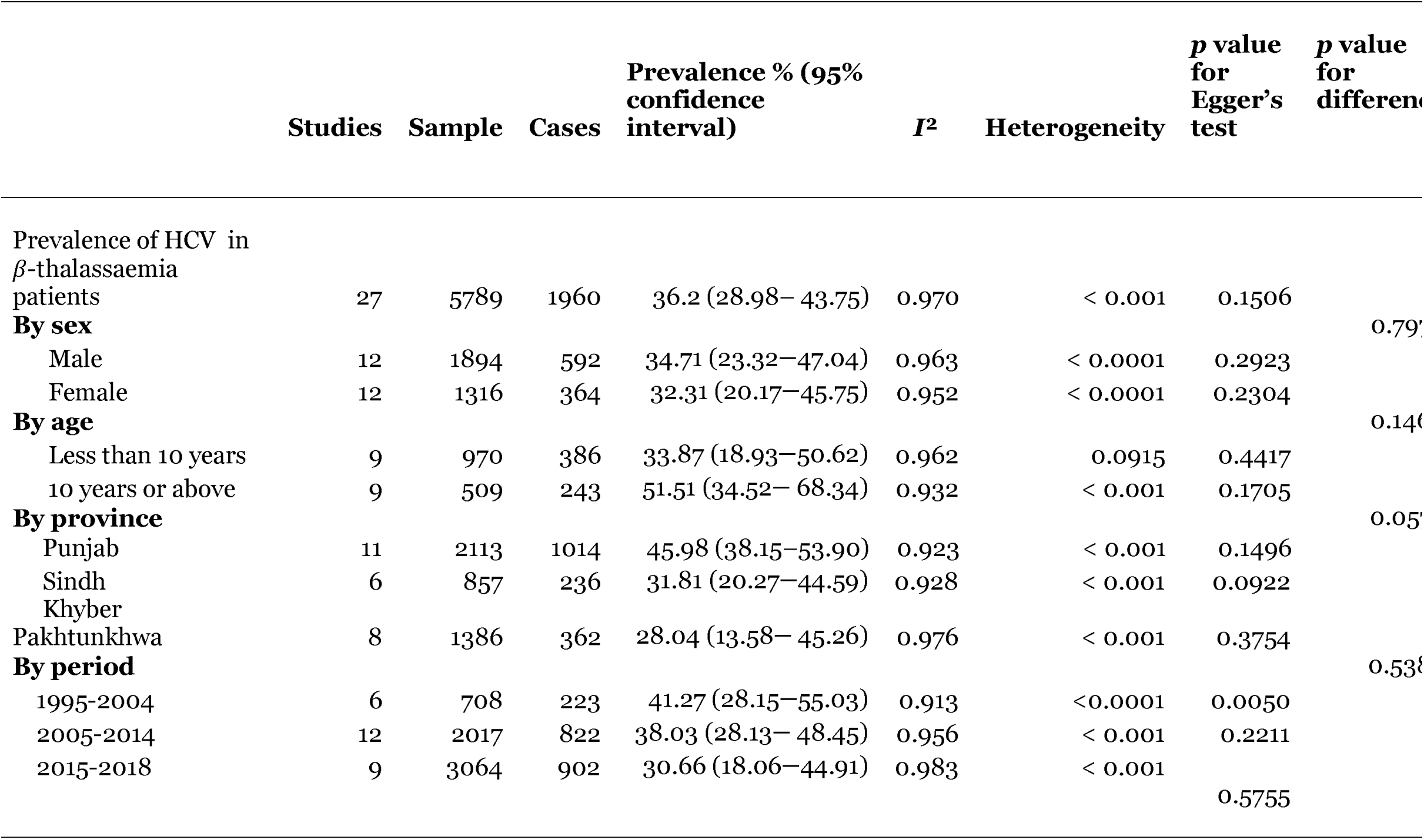
Summary statistics from meta-analyses of prevalence studies on HCV infection among thalassemia patients residing in Pakistan

**Figure 2:**
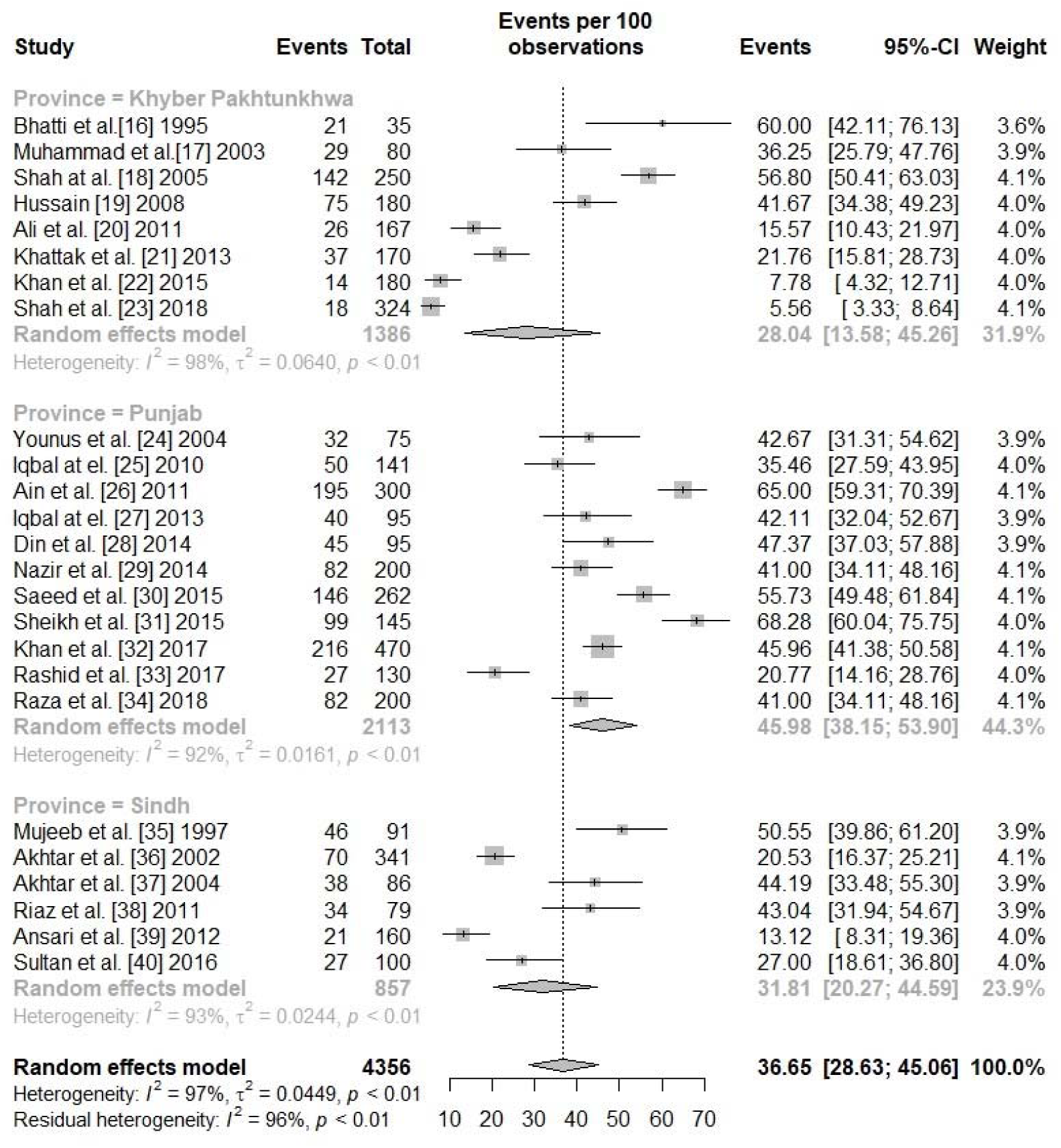
Forest plot of prevalence of HCV infection in *β*-thalassemia patients in Pakistan

**Figure 3:**
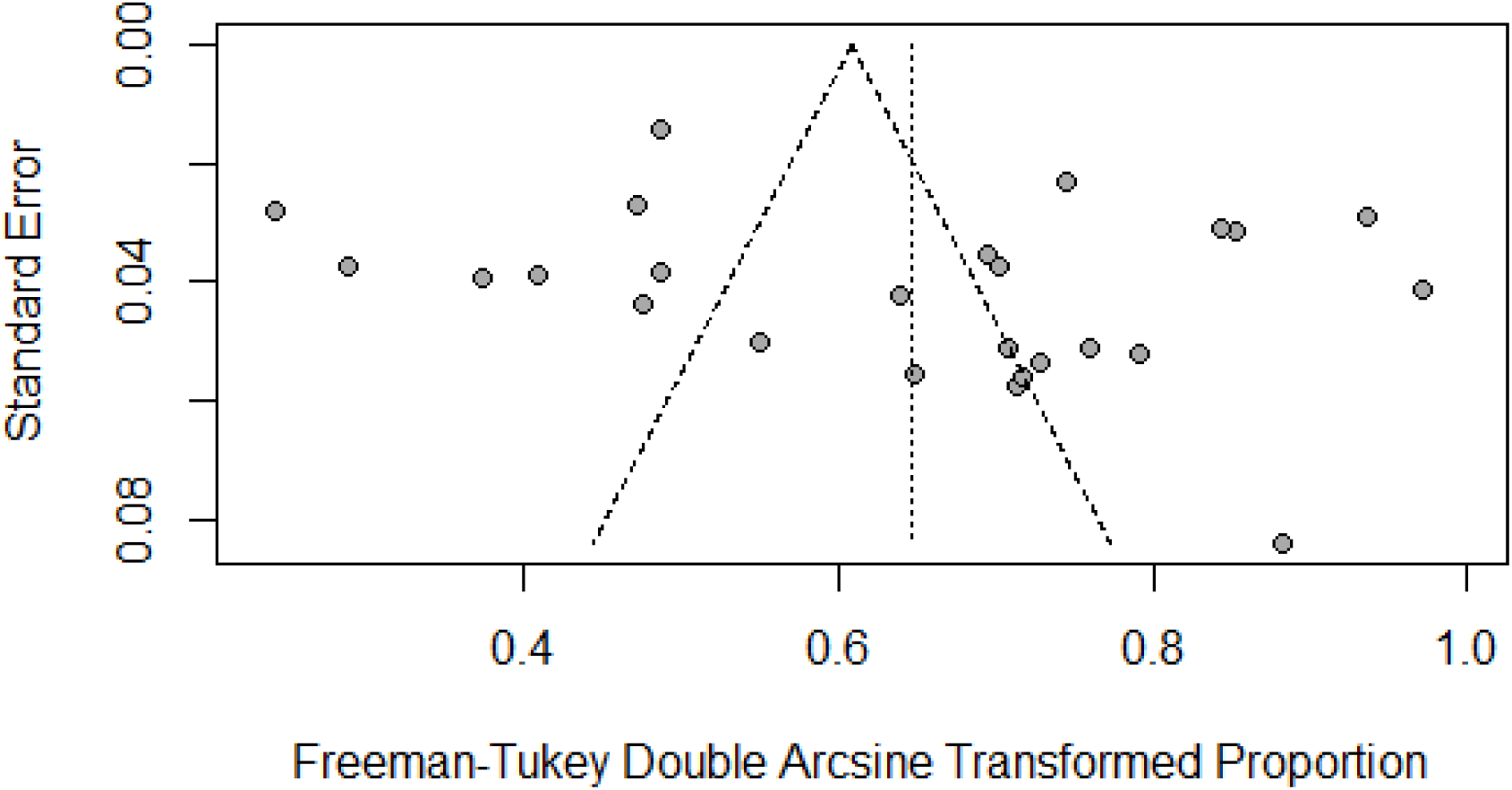
Funnel plot of the prevalence of HCV infection in *β*-thalassemia patients in Pakistan

The results of the univariable meta-regression analysis of the prevalence of HCV in *β*-thalassemia patients are presented in Table 3. The analysis shows that only geographical region (province) had a significant effect on the prevalence of HCV in *β*-thalassemia patients with a p-value < 0.1, while year of publication, year of data collection, sample size, proportion of males, average age of thalassemia patients had no significant effect on the observed HCV prevalence in *β*-thalassemia patients.

**Table 3.** Results of bivariate meta-regression for prevalence of HCV infection in thalassemia patients in Pakistan

| Covariate | Category | Number of Studies | Meta-regression Coefficient (%) | *p*-value | Variance explained *R*<sup>2</sup> (%) |
| --- | --- | --- | --- | --- | --- |
| Geographical region (Province) | Khyber Pakhtunkhwa | 8 | 1 |  |  |
|  | Punjab | 11 | 0.1873 | 0.0306 | 26.97 |
|  | Sindh | 6 | 0.0447 | 0.6582 |  |
| Year of publication |  | 27 | -0.0098 | 0.1176 | 1.33 |
| Year of data collection |  | 21 | -0.0076 | 0.3058 | 0.00 |
| Sample size | < 100 | 8 | 1 |  |  |
|  | >=100 | 19 | -0.1329 | 0.1187 | 2.92 |
| Proportion of males |  | 23 | -0.0003 | 0.9189 | 0.00 |
| Average age of patients |  | 25 | -0.0058 | 0.6598 | 3.41 |
| Sample size, continuous |  | 27 | -0.0098 | 0.1176 | 1.33 |

## Discussion

The aim of this study was to summarize the available literature on the prevalence of HCV infection among *β*-thalassemia patients and its associated risk factors in Pakistan. The result of the systematic review and meta-analysis showed that the pooled prevalence based on 27 studies was 36.21%— one in every three *β*-thalassemia patients in Pakistan have already been exposed to HCV infection. The prevalence of HCV among *β*-thalassemia patients, as revealed by this study is six times higher (36.21%) than in the general Pakistani population which is 6.2%^44^. In Pakistan, many patients with *β*-thalassemia have limited access to regular and safe blood transfusions. Possible reasons for this are the lack of altruistic voluntary blood donors and the inadequate testing of blood donations for HCV. Many blood transfusion centers and hospitals have inadequate resources and kits for screening blood donations.^5^ The root cause of the high prevalence is predominantly the lack of adequate regulation of blood banks and monitoring to assess compliance with transfusion safety standards. It is well recognized that, with proper regulation driven by policy makers, transfusion transmitted infections are markedly reduced^5^. Pakistan is a low resource country: the prevalence of HCV in *β*-thalassemia patients in Pakistan is higher than that in Iran^46^ (19%) or Bangladesh^47^ (14.7%). The findings of this study should act as a major safety alert for decision and policy-makers in the Pakistani health sector.

HCV infection prevalence among the *β*-thalassemia patients was observed across Pakistan in all provinces except Baluchistan and Gilgit-Baltistan, as we did not find any studies for these provinces. Our results showed that the prevalence of HCV among *β*-thalassemia patients was higher in Punjab (45.98%) than in Sindh (31.81%) and Khyber Pakhtunkhwa (28.04%).

In this paper, we found that the prevalence of HCV among *β*-thalassemia patients rises with age, increasing from 33.87% in the under 10 years age group compared to 51.51% in the 10 years or above age group. This is undoubtedly partly due to cumulative exposure to blood transfusions over a life time. Conversely, one could look at this more positively and suggest that the frequency of testing for HCV positive blood donations has improved and hence younger patients have a lower infection rate than their older fellow patients did when they were the same age, due to safer blood donations.

Furthermore, meta-regression analyses showed that there was no significant change in the prevalence of HCV among *β*-thalassemia patients over the past three decades (with both years of publication and year of data collection). The average age of the *β*-thalassemia patients was insignificant to the prevalence of HCV. Rather than age, it is the number of blood transfusions that play a vital role in the prevalence of HCV^42^, and we do not have data on this.

To the best of our knowledge, this paper is the first systematic review and meta-analysis to summarize the current data on the prevalence of HCV infection among *β*-thalassemia patients in Pakistan. The main strengths of this review are the use of a predefined and a comprehensive literature search strategy, and the involvement of two independent investigators in the whole review process and data extraction. No publication bias was found within our analyses which suggests that we are unlikely to have missed any significant studies that could change the findings. Furthermore, all articles which included in this meta-analysis had a low risk of bias in their methodological quality. As investigated by the meta-regression analysis, the methodological quality of the studies had no impact on pooled estimates. Three provinces of Pakistan were represented in the determination of HCV infection prevalence in *β*-thalassemia patients. On the other hand, the findings of this systematic review and meta-analysis have some limitations. Firstly, the meta-regression analysis was only based on bivariate analysis. We planned to perform a multivarible meta-regression analysis by considering all the factors simultaneously, however, it was not possible to use multivariable meta-regression analysis due to the small number of studies. A multivariable meta-regression analysis required at least ten studies per factor to estimate the meta-regression coefficients efficiently. Second and as is common in meta-analyses, our estimates revealed significant heterogeneity. It is possible that other sources of variation may have been missed in our analysis, such as the number of blood transfusions, type of *β*-thalassemia and genetic factors; but we were unable to test them due to data unavailability.

## Conclusions

The overall prevalence of hepatitis C virus among *β*-thalassemia patients in Pakistan was 36.21%, but varies from province to province. The prevalence is higher than in neighboring countries such as Iran and Bangladesh. Pakistan is a developing country and lacking in resources for appropriate blood screening facilities in thalassemia centers and hospitals. Lack of robust policies on transfusion safety as well as appropriate and rigorous monitoring of blood banks to ensure compliance with policies perpetuate the risk of transfusion transmitted infection with HCV. National and regional health programs should mandate and monitor the screening procedures so as to reduce the risk of transfusion transmitted infections such as HCV in the general population in *β*-thalassemia patients.

## Data Availability

All the data are inside the paper

## Data Availability

All the data are inside the paper

## Authors Contributions

SA, JAN did data collection and manuscript writing, conceived and designed

SA, JAN and AH did analysis & editing of manuscript

SA, FS and AH did review and final approval of manuscript

## Competing Interests

The authors declare no competing interests.

## Notes

### Competing Interest Statement

The authors have declared no competing interest.

